# Clinical Severity and mRNA Vaccine Effectiveness for Omicron, Delta, and Alpha SARS-CoV-2 Variants in the United States: A Prospective Observational Study

**DOI:** 10.1101/2022.02.06.22270558

**Authors:** Adam S. Lauring, Mark W. Tenforde, James D. Chappell, Manjusha Gaglani, Adit A. Ginde, Tresa McNeal, Shekhar Ghamande, David J. Douin, H. Keipp Talbot, Jonathan D. Casey, Nicholas M. Mohr, Anne Zepeski, Nathan I. Shapiro, Kevin W. Gibbs, D. Clark Files, David N. Hager, Arber Shehu, Matthew E. Prekker, Heidi L. Erickson, Matthew C. Exline, Michelle N. Gong, Amira Mohamed, Nicholas J. Johnson, Vasisht Srinivasan, Jay S. Steingrub, Ithan D. Peltan, Samuel M. Brown, Emily T. Martin, Arnold S. Monto, Akram Khan, Catherine L. Hough, Laurence W. Busse, Caitlin C. ten Lohuis, Abhijit Duggal, Jennifer G. Wilson, Alexandra June Gordon, Nida Qadir, Steven Y. Chang, Christopher Mallow, Carolina Rivas, Hilary M. Babcock, Jennie H. Kwon, Natasha Halasa, Carlos G. Grijalva, Todd W. Rice, William B. Stubblefield, Adrienne Baughman, Kelsey N. Womack, Jillian P. Rhoads, Christopher J. Lindsell, Kimberly W. Hart, Yuwei Zhu, Katherine Adams, Stephanie J. Schrag, Samantha M. Olson, Miwako Kobayashi, Jennifer R. Verani, Manish M. Patel, Wesley H. Self, For the Influenza and Other Viruses in the Acutely Ill (IVY) Network

## Abstract

**Objectives:** To characterize the clinical severity of COVID-19 caused by Omicron, Delta, and Alpha SARS-CoV-2 variants among hospitalized adults and to compare the effectiveness of mRNA COVID-19 vaccines to prevent hospitalizations caused by each variant.

**Design:** A case-control study of 11,690 hospitalized adults.

**Setting:** Twenty-one hospitals across the United States.

**Participants:** This study included 5728 cases hospitalized with COVID-19 and 5962 controls hospitalized without COVID-19. Cases were classified into SARS-CoV-2 variant groups based on viral whole genome sequencing, and if sequencing did not reveal a lineage, by the predominant circulating variant at the time of hospital admission: Alpha (March 11 to July 3, 2021), Delta (July 4 to December 25, 2021), and Omicron (December 26, 2021 to January 14, 2022).

**Main Outcome Measures:** Vaccine effectiveness was calculated using a test-negative design for COVID-19 mRNA vaccines to prevent COVID-19 hospitalizations by each variant (Alpha, Delta, Omicron). Among hospitalized patients with COVID-19, disease severity on the WHO Clinical Progression Ordinal Scale was compared among variants using proportional odds regression.

**Results:** Vaccine effectiveness of the mRNA vaccines to prevent COVID-19-associated hospitalizations included: 85% (95% CI: 82 to 88%) for 2 vaccine doses against Alpha; 85% (95% CI: 83 to 87%) for 2 doses against Delta; 94% (95% CI: 92 to 95%) for 3 doses against Delta; 65% (95% CI: 51 to 75%) for 2 doses against Omicron; and 86% (95% CI: 77 to 91%) for 3 doses against Omicron. Among hospitalized unvaccinated COVID-19 patients, severity on the WHO Clinical Progression Scale was higher for Delta than Alpha (adjusted proportional odds ratio [aPOR] 1.28, 95% CI: 1.11 to 1.46), and lower for Omicron than Delta (aPOR 0.61, 95% CI: 0.49 to 0.77). Compared to unvaccinated cases, severity was lower for vaccinated cases for each variant, including Alpha (aPOR 0.33, 95% CI: 0.23 to 0.49), Delta (aPOR 0.44, 95% CI: 0.37 to 0.51), and Omicron (aPOR 0.61, 95% CI: 0.44 to 0.85).

**Conclusions:** mRNA vaccines were highly effective in preventing COVID-19-associated hospitalizations from Alpha, Delta, and Omicron variants, but three vaccine doses were required to achieve protection against Omicron similar to the protection that two doses provided against Delta and Alpha. Among adults hospitalized with COVID-19, Omicron caused less severe disease than Delta, but still resulted in substantial morbidity and mortality. Vaccinated patients hospitalized with COVID-19 had significantly lower disease severity than unvaccinated patients for all the variants.

## INTRODUCTION

The coronavirus disease 2019 (COVID-19) pandemic has been defined by both the distribution of highly effective vaccines and the serial emergence of new severe acute respiratory syndrome coronavirus-2 (SARS-CoV-2) genetic variants [1]. Variants of concern are new genetic versions of the virus with increased transmissibility, a change in virulence or disease presentation, or a decrease in effectiveness of mitigation measures, available vaccines, or therapeutics [2]. There are now five WHO designated SARS-CoV-2 variants of concern: Alpha (B.1.1.7 and descendant lineages), Beta (B.1.351), Gamma (P.1), Delta (B.1.617.2 and AY lineages), and Omicron (B.1.1.529 and BA lineages).

First identified in Spring 2021, the highly contagious Delta variant rapidly replaced other SARS-CoV-2 variants and achieved global dominance by Summer 2021 [3]. Early studies suggested potential for increased risk of hospitalization for Delta-infected individuals compared to prior variants [4–6]. The highly divergent Omicron variant was identified in mid-November 2021 and quickly became the dominant variant in much of Europe and North America by late-December 2021 [7]. The overall risk of hospitalization among those infected with the Omicron variant appears to be lower than those infected with the Delta variant [8]. However, hospitalizations for Omicron infection do occur and disease severity and risk for progression to critical illness remain incompletely understood for this variant.

Understanding the epidemiology SARS-CoV-2 variants and the effectiveness of existing vaccines against them are essential to guide vaccination policies and development of new vaccines. Early studies suggested reduced vaccine effectiveness against infection and hospitalization for Omicron compared to earlier variants [9–11]. In most cases, estimates of vaccine effectiveness against the Omicron variant were based on cases that occurred during time periods in which the Omicron variant exceeded 50% in genomic surveillance. While efficient, these approaches have the potential for variant misclassification and inaccurate vaccine effectiveness estimates. Furthermore, little is known about the effectiveness of vaccines for the prevention of the most severe manifestations of COVID-19, including respiratory failure and death, for patients with Omicron infection.

Using observational study designs, the Influenza and Other Viruses in the Acutely Ill (IVY) Network in collaboration with the United States Centers for Disease Control and Prevention (CDC) is studying the effectiveness of COVID-19 vaccines against severe disease (Table S1) [12–15]. Here, we compare the clinical severity of COVID-19 caused by the SARS-CoV-2 Alpha, Delta and Omicron variants among hospitalized adults in the United States and the effectiveness of mRNA COVID-19 vaccines against each of these variants.

## METHODS

### Design and Setting

We conducted a prospective observational study at 21 hospitals in the United States, with enrollment of adults hospitalized with laboratory-confirmed COVID-19 and concurrent controls without COVID-19. A test-negative design was utilized to assess vaccine effectiveness [16]. This program was conducted by the IVY Network, which is a group consisting of geographically dispersed academic medical centers in the United States, coordinated from Vanderbilt University Medical Center, and funded by CDC (Table S1). Participants enrolled in the IVY program with hospital admission dates between March 11, 2021, and January 14, 2022 were included in this analysis. This program was approved as a public health surveillance activity with waiver of informed consent by CDC and all participating sites.

This analysis compared the Alpha, Delta, and Omicron SARS-CoV-2 variants in three ways: (1) vaccine effectiveness of the COVID-19 mRNA vaccines to prevent hospitalizations due to each variant; (2) disease severity among unvaccinated and vaccinated patients hospitalized with each variant; and (3) vaccine effectiveness of the COVID-19 mRNA vaccines to prevent disease progression to invasive mechanical ventilation or death after hospitalization with each variant.

### Participants

Sites prospectively screened hospitalized adults ≥18 years old for potential eligibility through daily review of hospital admission logs and electronic medical records. COVID-19 cases included those hospitalized with a clinical syndrome consistent with acute COVID-19 (≥1 of the following: fever; cough; shortness of breath; loss of taste; loss of smell; use of respiratory support for the acute illness; or new pulmonary findings on chest imaging consistent with pneumonia) and a positive molecular or antigen test for SARS-CoV-2 within 10 days of symptom onset. As described below, COVID-19 case patients were subclassified based on SARS-CoV-2 variant. Additionally, two control groups were enrolled: 1) “test-negative” controls were adults hospitalized with signs or symptoms consistent with acute COVID-19 who tested negative for SARS-CoV-2; and 2) “syndrome-negative” controls were adults hospitalized without signs or symptoms consistent with acute COVID-19 and who tested negative for SARS-CoV-2 [17]. Controls were selected from lists of eligible participants hospitalized within 2 weeks of enrollment of COVID-19 cases. Sites attempted to capture all COVID-19 cases admitted to the hospital during the surveillance period and targeted a case: control ratio of approximately 1:1. Cases and controls were not individually matched. Respiratory samples from participants were tested for SARS-CoV-2 both locally in clinical laboratories and centrally at a research laboratory (see Laboratory Analysis section). Cases tested positive for SARS-CoV-2 at a local laboratory, the central laboratory, or both, while controls tested negative for SARS-CoV-2 by all testing. Additional details about eligibility criteria and enrollment practices are described in Supplementary Appendix B [13,14].

### Data Collection

Trained personnel collected data on demographics, medical conditions, COVID-19 vaccination, and hospital course through participant (or proxy) interviews and standardized medical record review. Details of COVID-19 vaccination, including dates and location of vaccination, vaccine product, and lot number, were collected through a systematic process that included participant (or proxy) interview and source verification by vaccination card, hospital records, state vaccine registries, and vaccine records requested from clinics and pharmacies [13,14].

### Vaccination Status

Vaccine doses were classified as administered if source documentation of the dose was identified or if the participant/proxy reported a vaccine dose with a complete and plausible date and location. This analysis focused on COVID-19 mRNA vaccines authorized or approved for use in the United States, including BNT162b2 (Pfizer-BioNTech) and mRNA-1273 (Moderna). Participants were classified based on the number of mRNA vaccine doses received before illness onset: 0 doses (unvaccinated); 1 dose ≥14 days before illness (partially vaccinated); 2 doses ≥14 days before illness (fully vaccinated); or 3 doses ≥7 days before illness (boosted if immunocompetent, or with primary 3-dose series completed if immunocompromised). In the primary analysis, vaccine effectiveness was calculated for 2 vaccine doses for participants enrolled throughout the surveillance period and for 3 vaccine doses for participants enrolled after third doses were authorized in the United States [18,19]. In a secondary analysis, vaccine effectiveness was calculated for partial vaccination. Participants were excluded from this analysis if they received a COVID-19 vaccine other than an mRNA vaccine (e.g., the Ad26.COV2 vaccine from Janssen), more than 3 vaccine doses, or a third vaccine dose before they were authorized in the United States [18–20].

### Laboratory Analysis

Upper respiratory specimens (nasal swabs or saliva) were collected from participants, frozen, and shipped to Vanderbilt University Medical Center (Nashville, Tennessee), where they underwent reverse-transcription quantitative polymerase chain reaction (RT-qPCR) for detection of two SARS-CoV-2 nucleocapsid gene targets (N1 and N2) [21]. Respiratory specimens positive for SARS-CoV-2 were shipped to the University of Michigan (Ann Arbor, Michigan) for viral whole-genome sequencing using the ARTIC Network protocol on an Oxford Nanopore Technologies GridION instrument [22]. SARS-CoV-2 lineages were assigned using Pangolin [23]. The WHO variant assignment was as follows: Alpha (B.1.1.7), Beta (B.1.351), Gamma (P.1), Delta (B.1.617.2 and AY lineages), Omicron (B.1.1.529 or BA lineages).

### COVID-19 disease severity

We classified COVID-19 disease severity based on the highest severity state reached during the index COVID-19 hospital admission using a modified version of the WHO COVID-19 Clinical Progression Scale (Table S2) [13,24]. In this analysis of hospitalized patients, the scale levels included: hospitalized without supplemental oxygen (level 4), hospitalized with standard supplemental oxygen (level 5), hospitalized with high flow nasal cannula or non-invasive ventilation (level 6), hospitalized with invasive mechanical ventilation (level 7), hospitalized with mechanical ventilation and additional organ support (e.g., ECMO, vasopressors; level 8), and death (level 9). In addition to evaluating the full scale (levels 4-9) as an ordinal outcome, we also dichotomized the scale at level 7 to facilitate comparison between patients who experienced death or invasive mechanical ventilation (levels 7-9) vs those who did not experience death or invasive mechanical ventilation (levels 4-6).

### Statistical Analysis

COVID-19 cases were classified into Alpha, Delta, and Omicron categories using sequencing information for cases with lineages identified and by the predominant circulating variant at the time of hospital admission for those without a lineage identified. Periods of predominant circulation for Alpha, Delta and Omicron were defined based on time windows when each variant was identified in >50% of cases successfully sequenced in the study—Alpha period: March 11 – July 3, 2021; Delta period: July 4 – December 25, 2021; and Omicron period: December 26, 2021 – January 14, 2022. For analyses evaluating vaccine effectiveness to prevent hospitalization, evaluating cases and controls enrolled during the same time period was important to maintain accuracy of vaccine effectiveness estimates. For analyses evaluating vaccine effectiveness against hospitalization, cases and controls in each period (Alpha, Delta, and Omicron) were compared; cases were excluded from these analyses if they had a lineage identified by sequencing that was discordant with the period (for example, a Delta lineage identified in the Omicron period). For severity analyses, only cases were analyzed and maintaining a temporal relationship with a control group was not necessary; therefore, all cases with sequencing-confirmed Alpha, Delta, or Omicron lineage were analyzed regardless of admission date; in these analyses, variant group was classified by sequencing confirmation of Alpha, Delta or Omicron lineage, and then for other cases, by period.

Vaccine effectiveness of COVID-19 mRNA vaccines (BNT162b2 or mRNA-1273) to prevent hospitalization for COVID-19 was calculated using a test-negative design, in which the odds of antecedent vaccination were compared between cases and controls. Participants in the test-negative and syndrome-negative control groups were pooled based on analyses demonstrating highly similar vaccine coverage in the two control groups and nearly identical vaccine effectiveness estimates when either control group was used individually [14]. A multivariable unconditional logistic regression model was constructed with case-control status as the dependent variable, vaccination status (vaccinated vs. unvaccinated) as the primary independent variable and the following covariables selected *a priori* as potential confounders: calendar date of admission in biweekly intervals, US Department of Health and Human Services region (10 regions), age, sex, and self-reported race and Hispanic ethnicity. Post-hoc, the following variables were considered for potential inclusion as covariates but none of them changed the adjusted odds ratio (aOR) by more than 5% and were not included in the final analysis: number of comorbidities, smoking status, living in a long-term care facility before hospital admission and working in a healthcare setting. Vaccine effectiveness to prevent COVID-19 hospitalization [VE(hospitalization)] was calculated with the adjusted odds ratio (aOR) from this model as: VE(hospitalization) = (1 – aOR) × 100. Using this method, vaccine effectiveness against COVID-19 hospitalization as calculated separately for the Alpha, Delta, and Omicron. Vaccine effectiveness for two vaccine doses was calculated for each period, and for three vaccine doses for the Delta and Omicron periods. Within each period, vaccine effectiveness was also calculated for subgroups defined by: immunocompromised status [14]; age group (18-64 years; ≥65 years); burden of chronic medical conditions (0; ≥1 medical conditions); vaccine product (BNT162b2; mRNA-1273); and for two vaccine doses recipients, the time between the second vaccine dose and symptom onset (14-150 days; >150 days). This threshold of 150 days was selected based on the recommendation for a third (booster) dose of mRNA vaccine after 5 months for immunocompetent adults [23]. Results for subgroup analyses that had >150 cases and controls were reported.

COVID-19 severity by variant and by vaccination status was displayed by plotting the highest severity level on the modified WHO Clinical Progression Scale attained for each case. COVID-19 severity was compared among unvaccinated cases by variant (Alpha, Delta, Omicron) and between unvaccinated and vaccinated cases within each group. In these analyses of severity, patients vaccinated with either 2 or 3 doses of an mRNA vaccine were considered fully vaccinated. These calculations were performed using a multivariable proportional odds regression model with WHO ordinal scale as the dependent variable (levels 4-9), variant group (Alpha, Delta, Omicron) or vaccination status (unvaccinated, vaccinated) as the primary independent variable and the following covariables: age, sex, race and Hispanic ethnicity, and number of underlying medical conditions (0, 1, 2, 3, or ≥4 classes of chronic conditions). An adjusted proportional odds ratio (aPOR) >1.0 from these models indicated more severe disease for the later variant than a comparator earlier variant, for example Delta compared with Alpha, and Omicron compared with Delta.

Next, the vaccine effectiveness was calculated for mRNA vaccines to prevent disease progression to invasive mechanical ventilation or death among adults hospitalized with COVID-19. A multivariable logistic regression model was constructed with the composite of invasive mechanical ventilation or death as the dependent variable, vaccination status (vaccinated with 2 or 3 doses vs unvaccinated) as the primary independent variable and the same covariables as included in the severity proportional odds model. Vaccine effectiveness to prevent in-hospital disease progression was calculated as: VE(progression) = (1 – aOR) x 100. Using this method, vaccine effectiveness against disease progression was calculated separately for the Alpha, Delta, and Omicron groups.

Results were considered statistically significant if 95% confidence intervals for odds ratios did not include the null (OR=1.0) or two-sided p-values were <0.05. Missing values were not imputed; results were presented with denominators to indicate sample size in each analysis and models included participants with complete data for all variables in the model. Statistical analyses were performed with Stata Version 16 (College Station, TX) and SAS 9.4 (Cary, NC).

## RESULTS

### Participants and SARS-CoV-2 Variants

Between March 11, 2021 and January 14, 2022, 14,128 patients were enrolled across 21 hospitals; 2,438 patients were excluded from the primary analyses, most commonly for receiving >1 mRNA vaccine dose but not classifying into the two-dose or three-dose vaccine recipient categories (n=933) or for receiving a non-mRNA vaccine (n=682) (Figure S1). The population for analysis included 11,690 patients, including 5,728 COVID-19 cases and 5,962 controls.

SARS-CoV-2 sequencing results were obtained for 2,599/5,728 (45%) cases in the analytical population. Among cases with sequencing completed, during the Alpha period 242/421 (57%) cases had Alpha identified by sequencing, during the Delta period 1,867/1,930 (97%) cases had Delta identified by sequencing, and during the Omicron period 190/248 (77%) cases had Omicron identified by sequencing (Figure 1; Table S3).

**Figure 1.**
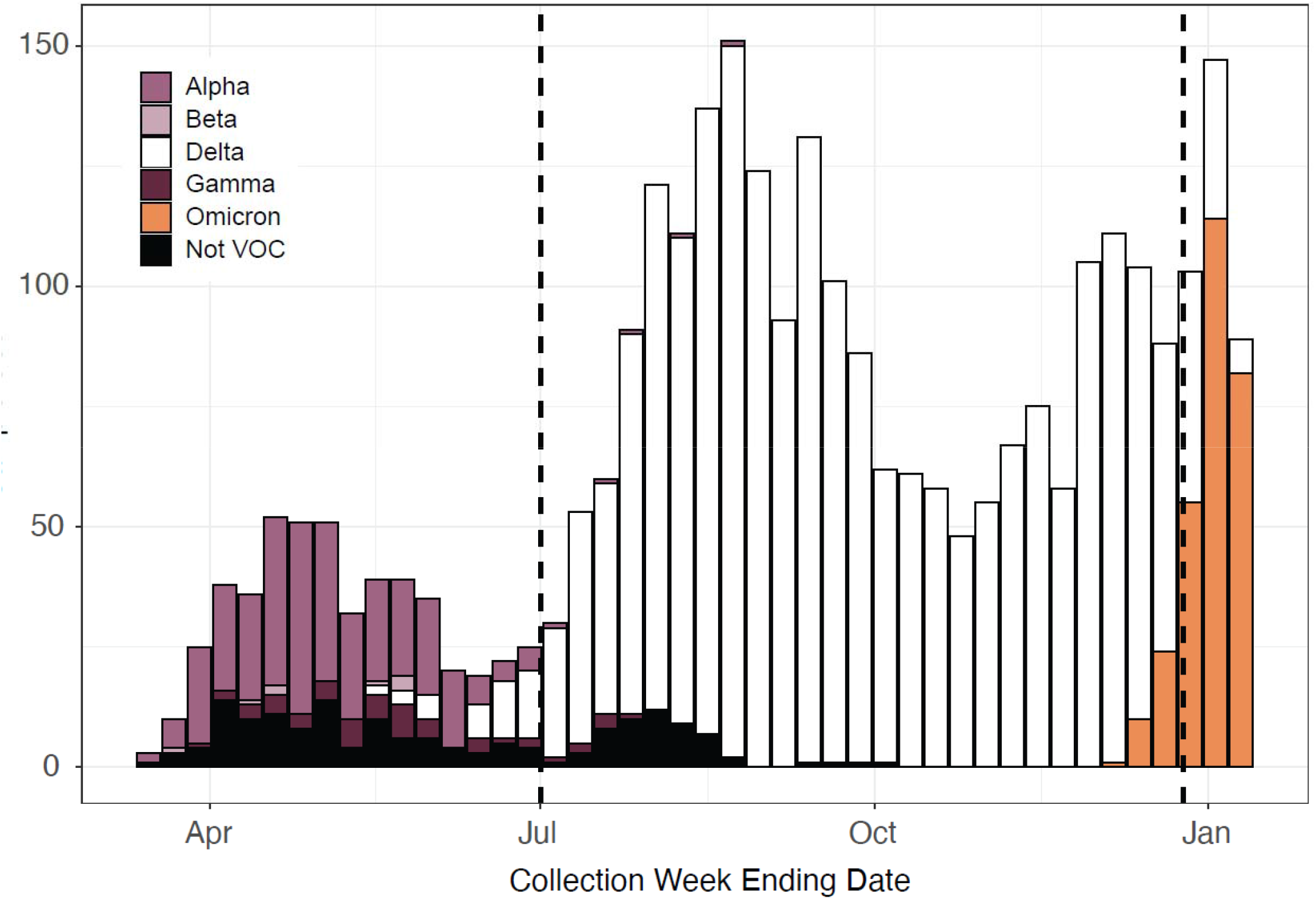
Sequenced SARS-CoV-2 variants by week among patients with COVID-19 enrolled in this study and hospitalized between March 11, 2021 and January 14, 2022 in 21 hospitals in the United States. Vertical dashed lines at July 4, 2021 and December 25, 2021 represent the start of the Delta period and Omicron period, respectively. This figure includes all cases enrolled in the program with a sequencing result, without restriction to cases included in the vaccine effectiveness analyses. SARS-CoV-2 variant lineages were identified for 3017 cases, including alpha (299), beta (8), delta (2209), gamma (52), omicron (286), and lineage not designated as variant of concern (163).

### Vaccine Effectiveness to Prevent COVID-19 Hospitalizations

After excluding 146/5,728 (3%) cases from the vaccine effectiveness against hospitalization analysis who had sequence-confirmed lineage discordant from the variant-predominant period (e.g., cases with sequencing-confirmed Delta variant during the Alpha or Omicron period), 5,582 cases and 5,962 controls were included in this part of the analysis. Cases included 1,072 from the Alpha period, 3,951 from the Delta period, and 559 from the Omicron period. Compared to cases in the Alpha and Delta period, cases in the Omicron group tended to be older, have more underlying medical conditions, and more likely to have ≥1 prior hospitalization in the past year (Table 1, Table S4). Consistent with increasing vaccine coverage in the United States population over time, a greater proportion of cases were vaccinated (2 or 3 doses of an mRNA vaccine) during the Omicron period (291/559, 52%) than the Alpha (119/1072, 11%) and Delta (1080/3951, 27%) periods.

**Table 1.** Characteristics of patients included in the evaluation of vaccine effectiveness to preventing COVID-19 hospitalizations, including hospitalized patients without COVID-19 (controls) and hospitalized patients with COVID-19 (cases) during the Alpha period (March 11 - July 3, 2021), Delta period (July 4 - December 25, 2021), and Omicron period (December 26, 2021-January 14, 2022). (Baseline characteristics for cases limited to those with a sequencing-confirmed variant are shown in Table S4.)

| Patient Characteristics | All controls<br>(n=5962) | Alpha cases<br>(n=1072) | Delta cases<br>(n=3951) | Omicron cases<br>(n=559) |
| --- | --- | --- | --- | --- |
| Age in years, median (IQR) | 63 (50-72) | 56 (43-65.5) | 57 (43-69) | 62 (49-73) |
| Female sex, No. (%) | 2975 (49.9) | 519 (48.4) | 1803 (45.6) | 264 (47.2) |
| Race and ethnicity, No. (%) |  |  |  |  |
| Non-Hispanic White | 3611 (60.6) | 484 (45.1) | 2183 (55.3) | 279 (49.9) |
| Non-Hispanic Black | 1240 (20.8) | 285 (26.6) | 820 (20.8) | 127 (22.7) |
| Hispanic, any race | 772 (12.9) | 220 (20.5) | 695 (17.6) | 111 (19.9) |
| Non-Hispanic, Other | 253 (4.2) | 63 (5.9) | 179 (4.5) | 33 (5.9) |
| Unknown | 86 (1.4) | 20 (1.9) | 74 (1.9) | 9 (1.6) |
| US Census region, No. (%) |  |  |  |  |
| Northeast | 885 (14.8) | 158 (14.7) | 686 (17.4) | 159 (28.4) |
| South | 2371 (39.8) | 395 (36.8) | 1544 (39.1) | 196 (35.1) |
| Midwest | 1374 (23.0) | 248 (23.1) | 978 (24.8) | 104 (18.6) |
| West | 1332 (22.3) | 271 (25.3) | 743 (18.8) | 100 (17.9) |
| Resident of long-term care facility, No. / Total (%) | 321/5778 (5.6) | 25/1039 (2.4) | 120/3795 (3.2) | 30/534 (5.6) |
| ≥1 prior hospitalization in past year, No. / Total (%) | 3031/5537 (54.7) | 282/956 (29.5) | 1015/3682 (27.6) | 226/535 (42.2) |
| Current tobacco use, No. / Total (%) | 1016/5302 (19.2) | 103/887 (11.6) | 366/3451 (10.6) | 59/470 (12.6) |
| Number of chronic medical conditions‡, median (IQR) | 2 (1-3) | 1 (1-3) | 1 (0-3) | 2 (1-3) |
| Categories of medical conditions‡ |  |  |  |  |
| Chronic cardiovascular disease | 4158 (69.7) | 589 (54.9) | 2141 (54.2) | 359 (64.2) |
| Chronic pulmonary disease | 1973 (33.1) | 231 (21.5) | 827 (20.9) | 151 (27.0) |
| Diabetes mellitus | 1962 (32.9) | 316 (29.5) | 1135 (28.7) | 164 (29.3) |
| Immunocompromising condition* | 1458 (24.5) | 172 (16.0) | 659 (16.7) | 138 (24.7) |
| Obesity, No. / Total (%) | 2391/5900 (40.5) | 616/1056 (58.3) | 2099/3909 (53.7) | 260/556 (46.8) |
| Vaccination status |  |  |  |  |
| Unvaccinated | 2054 (34.5) | 953 (88.9) | 2871 (72.7) | 268 (47.9) |
| 2 doses (<150 days) | 2029 (34.0) | 119 (11.1) | 352 (8.9) | 34 (6.1) |
| 2 doses (≥150 days) | 1411 (23.7) | 0 (0) | 667 (16.9) | 177 (31.7) |
| 3 doses | 468 (7.8) | 0 (0) | 61 (1.5) | 80 (14.3) |
| If vaccinated, vaccine product received, No. / Total (%) |  |  |  |  |
| BNT162b2 (Pfizer-BioNTech) | 2269/3908 (58.1) | 81/119 (68.1) | 708/1080 | 203/291 (69.8) |
|  |  |  | (65.6) |  |
| mRNA-1273 (Moderna) | 1615/3908 (41.3) | 37/119 (31.1) | 368/1080 (34.1) | 84/291 (28.9) |
| Mixed products | 24/3908 (0.6) | 1/119 (0.8) | 4/1080 (0.4) | 4/291 (1.4) |
| Days since dose 3 if 3 doses received, median (IQR) | 41 (23-64) | --- | 38 (23-65) | 69.5 (41.5-97) |

Definitions: IQR = interquartile range; US = United States
\*Immunocompromising conditions were obtained using structured medical chart review and defined as 1 or more of the following: active solid organ cancer (active cancer defined as treatment for the cancer or newly diagnosed cancer in the past 6 months), active hematologic cancer, HIV infection without AIDS, AIDS, congenital immunodeficiency syndrome, previous splenectomy, previous solid organ transplant, immunosuppressive medication, systemic lupus erythematosus, rheumatoid arthritis, psoriasis, scleroderma, or inflammatory bowel disease, including Crohn's disease or ulcerative colitis.

Vaccine effectiveness for two doses of mRNA vaccine to prevent COVID-19 hospitalization was 85% (95% CI: 82 to 88%) in the Alpha period, 85% (95% CI: 83 to 87%) in the Delta period, and 65% (95% CI: 51 to 75%) in the Omicron period (Figure 2). Vaccine effectiveness for three mRNA vaccine doses in the Omicron period was 86% (95% CI: 77 to 91%), which was similar to the effectiveness of two doses during the Alpha and Delta periods. Within the Delta period, vaccine effectiveness for two vaccine doses was lower when the second vaccine dose was >150 days before illness onset (81%; 95% CI: 78 to 84%) than 14-150 days (88%; 95% CI: 86 to 90%). Within the Delta period, vaccine effectiveness of three vaccine doses (94%; 95% CI: 92 to 95%) was higher than two doses, with high vaccine effectiveness observed after a third dose in both immunocompetent (97%; 95% CI: 95 to 98%) and immunocompromised (87%; 95% CI: 78 to 92%) patients. Within each period, vaccine effectiveness was lower for immunocompromised patients compared to immunocompetent patients and lower for the BNT162b2 vaccine than the mRNA-1273 vaccine. Vaccine effectiveness results for partial vaccination (either 1 dose of an mRNA vaccine or 2 doses with the second dose received <14 days before illness onset) are described in Supplementary Appendix C.

**Figure 2.**
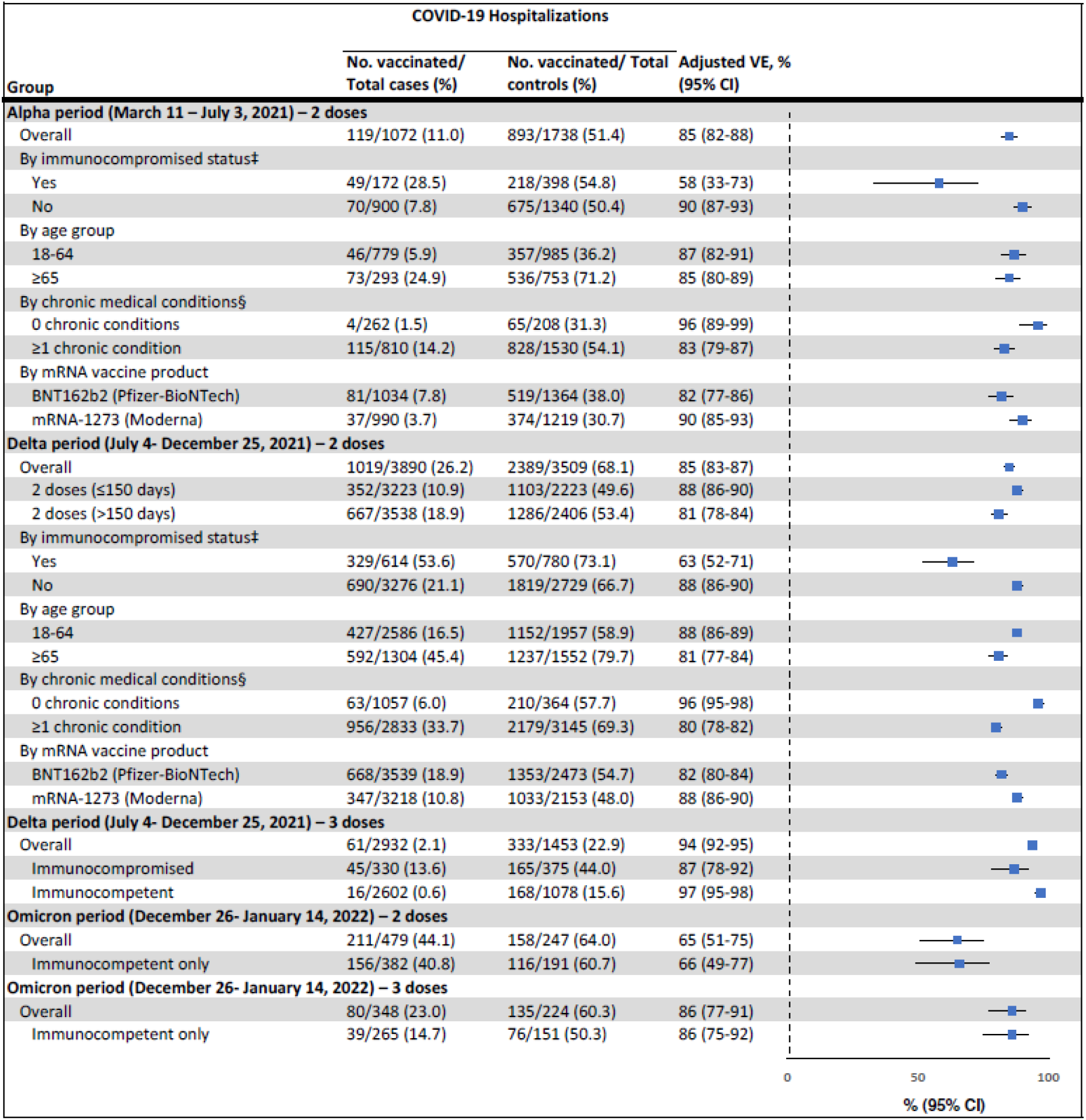
Vaccine effectiveness of COVID-19 mRNA vaccines to prevent COVID-19 hospitalizations by variant group, including Alpha, Delta, and Omicron. Definitions: VE = vaccine effectiveness ‡ Immunocompromising conditions were obtained using structured medical chart review and defined as 1 or more of the following: active solid organ cancer (active cancer defined as treatment for the cancer or newly diagnosed cancer in the past 6 months), active hematologic cancer, HIV infection without AIDS, AIDS, congenital immunodeficiency syndrome, previous splenectomy, previous solid organ transplant, immunosuppressive medication, systemic lupus erythematosus, rheumatoid arthritis, psoriasis, scleroderma, or inflammatory bowel disease, including Crohn’s disease or ulcerative colitis. § Chronic medical conditions were obtained using structured medical chart review and defined as conditions within 1 or more of the following categories: cardiovascular disease, neurologic disease, pulmonary disease, gastrointestinal disease, endocrine disease, renal disease, hematologic disease, malignancy, immunosuppression not captured in other categories, autoimmune condition, or other condition (sarcoidosis, amyloidosis, or unintentional weight loss ≥10 pounds (4.5 kg) in the last 90 days).

### COVID-19 Disease Severity

The severity analysis included data collected through January 31, 2022. Of 5,728 case patients in the study, 5,413 (95%) had complete clinical outcomes data and were included in the severity analysis, including 1,060 in the Alpha group, 3,788 in the Delta group, and 565 in the Omicron group. Overall, including both vaccinated and unvaccinated patients, 582/5,413 (11%) COVID-19 patients died within 28 days during the index hospitalization, including 81/1,060 (8%) in the Alpha group, 461/3,788 (12%) in the Delta group, and 40/565 (7%) in the Omicron group.

Among unvaccinated cases, COVID-19 severity on the WHO Clinical Progression Scale was highest for the Delta group (Delta vs Alpha aPOR 1.28, 95% CI: 1.11 to 1.46) and lowest for the Omicron group (Omicron vs Alpha aPOR 0.79, 95% CI: 0.62 to 1.01; Omicron vs Delta aPOR 0.61, 95% CI: 0.49 to 0.77) (Figure 3). Among unvaccinated cases, in-hospital death occurred in 76/944 (8%) in the Alpha group, 323/2,743 (12%) in the Delta group, and 25/272 (9%) in the Omicron group (Table 2). COVID-19 severity on the WHO Clinical Progression Scale was substantially lower for vaccinated cases than unvaccinated cases in each variant group, including Alpha (aPOR: 0.33; 95% CI: 0.23-0.49), Delta (aPOR 0.44; 95% CI: 0.37 to 0.51), and Omicron (aPOR: 0.61; 95% CI: 0.44 to 0.85).

**Table 2.** In-hospital clinical outcomes among adults hospitalized with COVID-19 by variant group (Alpha, Delta, Omicron).

| Outcome | Alpha Group (n=1060) |  |  |  | Delta Group (n=3788) |  |  |  | Omicron Group (n=565) |  |  |
| --- | --- | --- | --- | --- | --- | --- | --- | --- | --- | --- | --- |
|  | Vaccinated | Unvaccinated | P* |  | Vaccinated | Unvaccinated | P* |  | Vaccinated | Unvaccinated | P* |
| Death, no. (%) | 5/116 (4.3) | 76/944 (8.1) | 0.15 |  | 138/1045 (13.2) | 323/2743 (11.8) | 0.23 |  | 15/293 (5.1) | 25/272 (9.2) | 0.059 |
| Invasive mechanical ventilation, no. (%) | 7/116 (6.0) | 201/944 (21.3) | <0.001 |  | 152/1045 (14.5) | 681/2743 (24.8) | <0.001 |  | 35/293 (11.9) | 49/272 (18.0) | 0.043 |
| Composite of death or invasive mechanical ventilation, no. (%) | 10/116 (8.6) | 218/944 (23.1) | <0.001 |  | 210/1045 (20.1) | 748/2743 (27.3) | <0.001 |  | 42/293 (14.3) | 54/272 (19.9) | 0.08 |
| Admitted to intensive care unit, No. (%) | 24/116 (20.7) | 353/944 (37.4) | <0.001 |  | 321/1045 (30.7) | 1179/2742 (43.0) | <0.001 |  | 66/293 (22.5) | 89/271 (32.8) | 0.006 |
| Non-invasive ventilation, no. (%) | 15/116 (12.9) | 167/944 (17.7) | 0.20 |  | 151/1045 (14.4) | 470/2743 (17.1) | 0.046 |  | 39/293 (13.3) | 43/272 (15.8) | 0.40 |
| High-flow oxygen therapy, no. (%) | 15/116 (12.9) | 324/944 (34.3) | <0.001 |  | 289/1045 (27.7) | 1148/2743 (41.9) | <0.001 |  | 59/293 (20.1) | 84/272 (30.9) | 0.003 |
| Vasopressors, no. (%) | 5/116 (4.3) | 191/944 (20.2) | <0.001 |  | 155/1045 (14.8) | 647/2743 (23.6) | <0.001 |  | 36/293 (12.3) | 46/272 (16.9) | 0.12 |
| New renal replacement therapy, no. (%) | 6/116 (5.1) | 43/944 (4.6) | 0.91 |  | 49/1045 (4.7) | 159/2743 (5.8) | 0.18 |  | 14/293 (4.8) | 12/272 (4.4) | 0.84 |
| Hospital length among survivors, median (IQR) | 5 (3-8) | 5 (3-9) | 0.56 |  | 5 (3-9) | 6 (3-10) | <0.001 |  | 5 (3-9) | 6 (3-11) | 0.050 |
| Venous thromboembolic event, No. (%) | 7/116 (6.0) | 55/944 (5.8) | 0.93 |  | 46/1045 (4.4) | 250/2743 (9.1) | <0.001 |  | 15/293 (5.1) | 22/272 (8.1) | 0.15 |
| Stroke, No. (%) | 0/116 (0) | 18/944 (1.9) | 0.13 |  | 9/1045 (0.9) | 44/2743 (1.6) | 0.08 |  | 3/293 (1.0) | 4/272 (1.5) | 0.63 |
| Myocardial infarction, No. (%) | 1/116 (0.9) | 19/944 (2.0) | 0.39 |  | 29/1045 (2.8) | 58/2743 (2.1) | 0.23 |  | 5/293 (1.7) | 5/272 (1.8) | 0.91 |

\* P-values obtained using chi-square testing, not adjusting for other factors.

**Figure 3.**
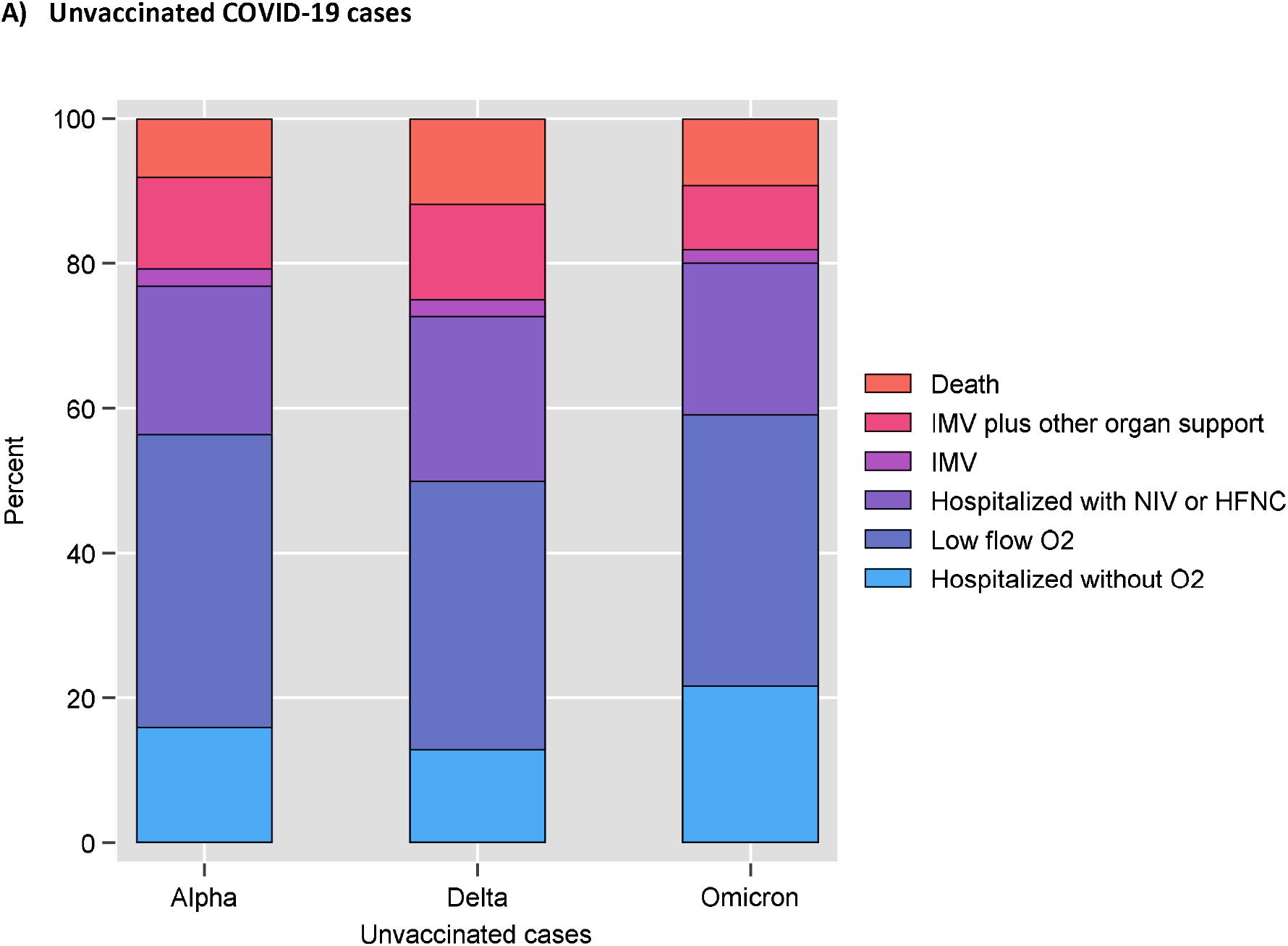
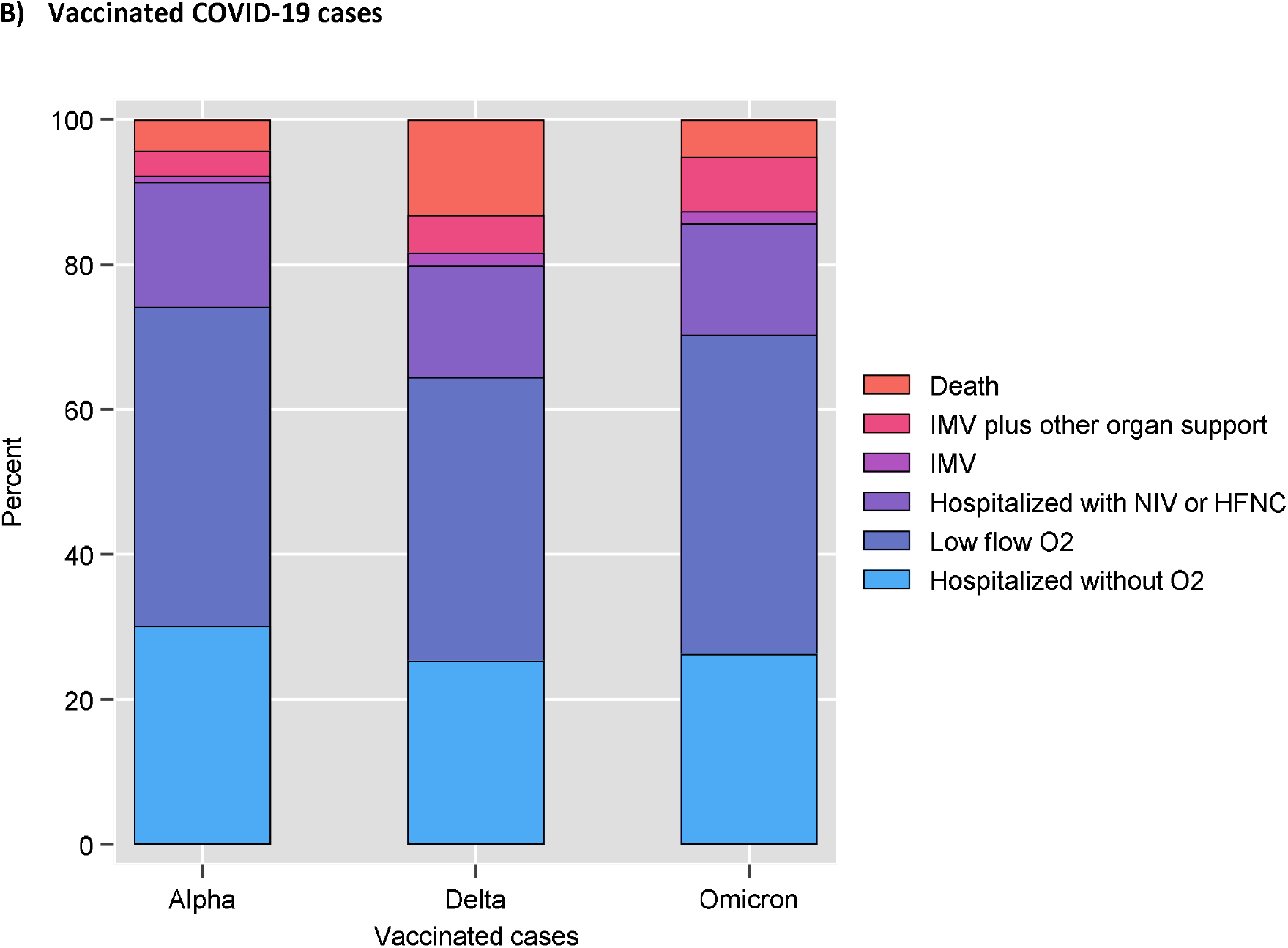
COVID-19 disease severity during index hospitalization among adults hospitalized with COVID-19, by SARS-CoV-2 variant for (A) unvaccinated patients, and (B) vaccinated patients. Disease severity was classified based on the highest severity level reached on the World Health Organization Clinical Progression Scale, which ranged from hospitalized without oxygen therapy (lowest level) to death (highest level). Among unvaccinated patients, severity was higher for Delta than Alpha (aPOR: 1.28, 95% CI: 1.11 to 1.46), lower for Omicron than Delta (aPOR: 0.61, 95% CI: 0.49 to 0.77). For each variant, severity was lower for vaccinated patients (2 or 3 doses of an mRNA vaccine) than unvaccinated patients, including for Alpha (aPOR: 0.33, 95% CI: 0.23 to 0.49), Delta (aPOR: 0.44, 95% CI: 0.37 to 0.51), and Omicron (aPOR: 0.61, 95% CI: 0.44 to 0.85). Data represented in the figures are shown in the accompanying table.

**Figure 3.**
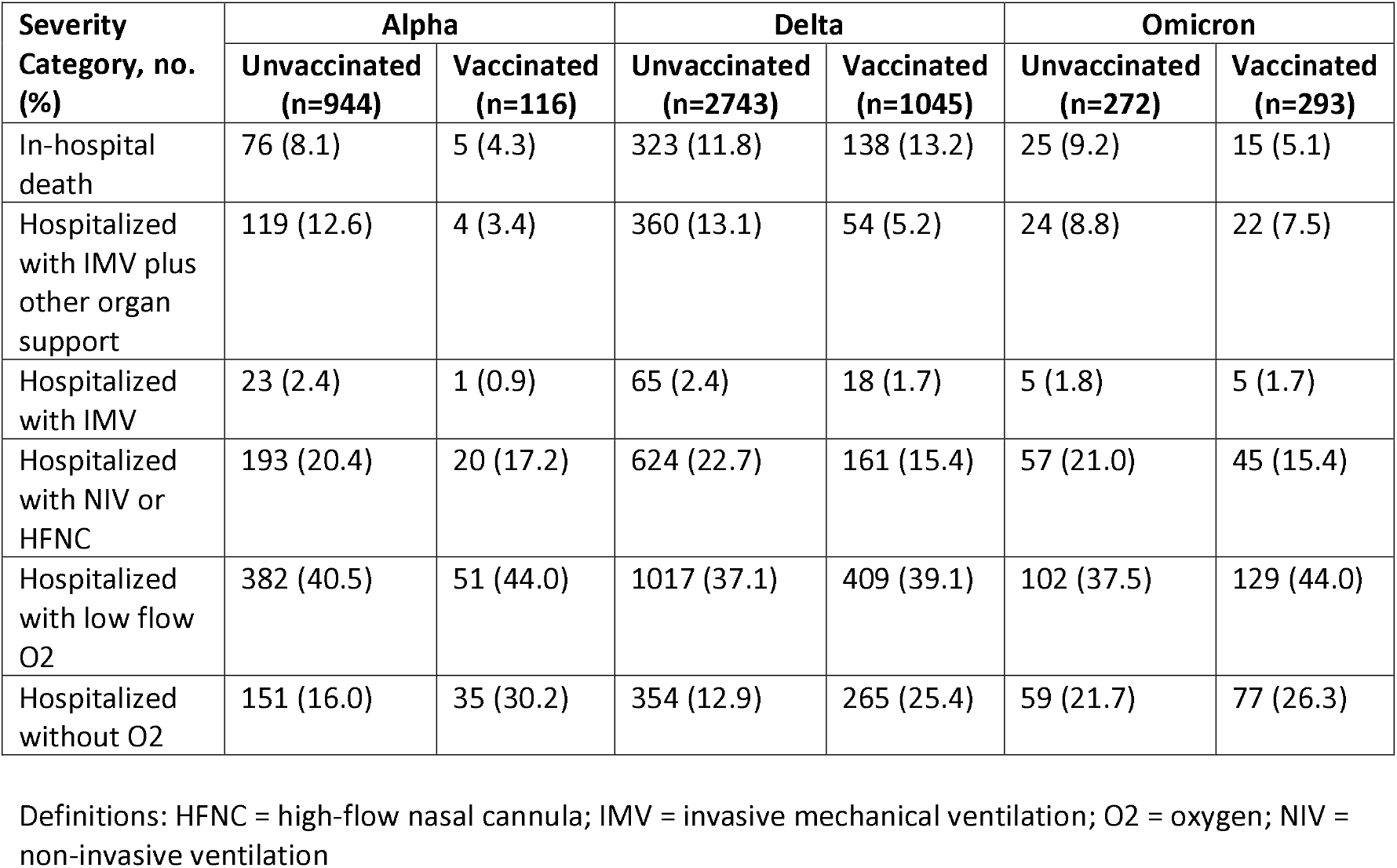
accompanying data table.

Across all variants, vaccinated COVID-19 patients who died tended to be old and had multiple medical conditions or immunocompromising conditions. Compared with the 424 unvaccinated COVID-19 patients who died, 158 vaccinated patients who died were older (median 72 vs 61 years; p<0.001), more likely to be immunocompromised (41% versus 13%; p<0.001), had more categories of chronic medical conditions (median 3 versus 2; p<0.001), and had more prescribed medications prior to hospital admission (median 10 versus 5; p<0.001).

### Vaccine Effectiveness to Prevent COVID-19 Disease Progression after Hospitalization

Among patients hospitalized with COVID-19, vaccine effectiveness of mRNA vaccination (2 or 3 doses) to prevent progression to invasive mechanical ventilation or death was 76% (95% CI: 53 to 88%) for Alpha, 44% (95% CI: 32 to 54%) for Delta, and 46% (95% CI: 12 to 67%) for Omicron. Vaccine effectiveness to prevent disease progression was observed for immunocompetent patients for all three variants, but not observed for immunocompromised patients for Delta or Omicron (Figure 4).

**Figure 4.**
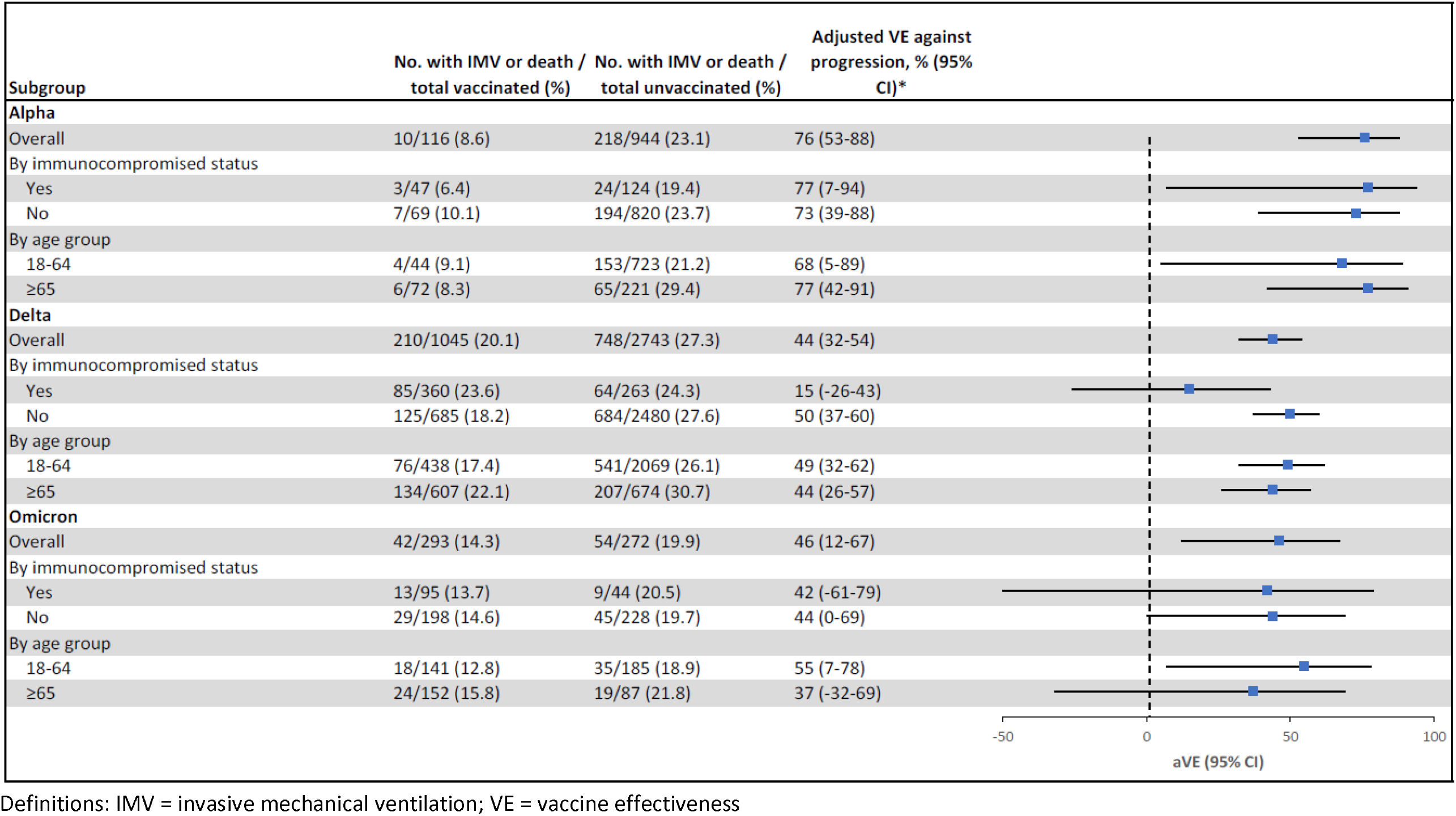
Vaccine effectiveness of two or three doses of COVID-19 mRNA vaccines among adults hospitalized with COVID-19 to prevent disease progression to invasive mechanical ventilation or death, by SARS-CoV-2 variant.

## DISCUSSION

### Principal Findings

The predominant circulating SARS-CoV-2 variant in the United States changed from Alpha to Delta in July 2021 and then to Omicron in December 2021. Understanding the disease severity caused by each variant and the effectiveness of available vaccines against them is essential for guiding vaccination policies and directing future vaccine development. The mRNA COVID-19 vaccines that were authorized for use in the United States in 2020 (BNT162b2 and mRNA-1273) were highly effective at preventing hospitalizations for all three variants during the subsequent year. However, three doses of an mRNA vaccine were necessary to achieve similar effectiveness against Omicron in the winter of 2021-2022 as two doses achieved for Alpha and Delta variants earlier in the year. Furthermore, while COVID-19 hospitalizations did occur among vaccinated patients, vaccination was associated with reduced risk of progression to invasive mechanical ventilation or death for all three variants.

Among unvaccinated hospitalized adults with COVID-19, Delta variant caused the most severe disease, followed by Alpha, and then Omicron. Among hospitalized unvaccinated patients, COVID-19 caused by Omicron was about 79% as severe as Alpha and 61% as severe as Delta. However, the Omicron variant did cause a substantial amount of critical illness and death, with 15% of all patients hospitalized with Omicron (vaccinated and unvaccinated) progressing to invasive mechanical ventilation and 7% dying in the hospital.

### Strengths and Limitations

This work has several strengths. The vaccine effectiveness analyses applied a test-negative design to a large, hospitalized population of patients with symptomatic, laboratory-confirmed COVID-19 and concurrent controls, which enabled control for healthcare seeking behavior, robust subgroup analyses, and evaluation of outcomes beyond hospital admission, including level of respiratory support and mortality. Ascertainment of vaccination status was robust, with trained personnel conducting patient interviews and searching multiple sources of vaccination records on a patient-by-patient basis. Respiratory samples collected in the study underwent centralized RT-PCR testing and viral whole genome sequencing, which enabled precise characterization of time periods dominated by different variants.

The study also had limitations. First, use of hospitalized controls might lead to biased estimates if control patients had different characteristics than people in the general community; however, vaccine coverage in the control population within this study tracked closely with that in the adult population in the United States [25], which lessens this potential concern. Second, this study only evaluated hospitalized patients and thus does not inform vaccine effectiveness against mild COVID-19 or differences in disease severity among SARS-CoV-2 variants in the outpatient setting. Third, the study only evaluated mRNA vaccines and did not assess other types of COVID-19 vaccines. Fourth, the analyses of in-hospital severity did not account for potential differences in clinical management during the Alpha, Delta, and Omicron periods that may have impacted outcomes. Fifth, while the test negative design is the preferred method for evaluating vaccine effectiveness with observational data [16], it has known potential limitations, including collider bias [26,27]; the risk of collider bias was minimized in the current study by evaluating only severely ill patients [16]. Sixth, sequencing did not identify a variant for some cases, typically those with low viral loads in the respiratory sample that underwent testing. Variant classification for cases without a sequencing-confirmed variant was based on the predominant circulating variant at the time; variant misclassification was possible for these cases.

### Comparison with Other Studies

Earlier studies from England [5] and Scotland [4] found an increased risk of hospital admission for Delta variant compared to Alpha variant. More recent studies have suggested that persons diagnosed with Omicron variant COVID-19 are less likely than those with Delta to be hospitalized [8]. This study adds robust measurements of disease severity after hospital admission and demonstrates Delta variant caused more severe disease than Alpha and Omicron variants, driven largely by higher rates of advanced respiratory support.

Emerging vaccine effectiveness estimates globally suggest reduced effectiveness against Omicron compared with prior variants [28–30], including an estimate of 70% vaccine effectiveness for two doses of the BNT162b2 vaccine to prevent Omicron hospitalizations in South Africa in November-December 2021 [9]. Using electronic health record data from sites across the United States, the VISION Network recently estimated mRNA vaccine effectiveness against Omicron hospitalizations to be to 52% for two vaccine doses with the second dose received within 180 days before illness onset, 38% for 2 doses received >180 days before illness onset, and 82% for 3 vaccine doses [11]. Prior studies largely relied on estimating predominant circulating SARS-CoV-2 variants from external data. This study adds vaccine effectiveness results against severe disease using sequence data from within the study and demonstrates strong protection against Omicron for three mRNA vaccine doses in the first several months after receipt of a third dose.

### Policy Implications

These data indicate that Omicron-variant COVID-19 is a serious disease among those who are hospitalized, and preventative measures are indicated. Vaccination with existing mRNA vaccine formulations is an effective preventative measure against Omicron, both for the prevention of hospitalization, and among those hospitalized, for the prevention of progression to critical illness and death. COVID-19 deaths in vaccinated individuals do occur, including with the Omicron variant, mostly in the elderly, the immunocompromised, or those with multiple medical comorbidities. These findings support recent recommendations in the United States for third mRNA vaccine doses for both immunocompetent [19] and immunocompromised [18] adults as a key approach to protecting populations against the Omicron variant.

The serial emergence of new SARS-CoV-2 variants, including Delta and Omicron, has challenged public health agencies to develop vaccine policies that counter the impact of waning immunity (the decline in protection of vaccine doses over time against the same variant) and viral immune evasion (new viral variants that are less susceptible to existing vaccines). Vaccine booster doses of the same vaccine formulation used in the primary vaccine series are designed to counter waning immunity. Significant viral immune evasion would require new vaccine formulations targeting new variants to maintain protection. Boosters were implemented in several countries in response to COVID-19 spikes with emergence of the Delta variant. This study suggests that these booster doses were largely effective in preventing severe disease with both Delta and the subsequent Omicron variant. As the COVID-19 pandemic continues to evolve, routine vaccine effectiveness monitoring, especially against severe disease, and surveillance programs to identify viral variants will be essential to inform decisions about booster vaccine policies and vaccine strain updates.

## Conclusions

In this large study of hospitalized adults in the United States, mRNA vaccines provided strong protection against COVID-19 hospitalization caused by the Alpha, Delta, and Omicron variants. Furthermore, vaccination reduced the risk for COVID-19 progressing to critical illness or death for each of the variants. While disease severity for patients hospitalized with COVID-19 was somewhat lower for Omicron than the Alpha and Delta variants, patients hospitalized with Omicron-variant COVID-19 still had a substantial risk for critical illness and death. These findings suggest that vaccination against COVID-19, including a third dose of an mRNA vaccine, is critical for protecting populations against COVID-19-associated morbidity and mortality.

## NOTES

### Contributions

Guarantors of this work include Dr. Self (protocol and data integrity), Dr. Tenforde (statistical analysis), Dr. Lauring (viral sequencing laboratory methods), and Dr. Chappell (RT-PCR laboratory methods). Contributions of each author include the following. Responsibility for decision to submit the manuscript: Self. Composed the initial manuscript draft: Lauring, Tenforde, Chappell, Patel, Self (the authors alone wrote the manuscript without outside assistance). Conceptualization of study methods: Lauring, Tenforde, Chappell, Talbot, Lindsell, Grijalva, Schrag, Kobayashi, Verani, Patel, Self. Data Collection: Lauring, Chappell, Gaglani, Ginde, McNeal, Ghamande, Douin, Talbot, Casey, Mohr, Zepeski, Shapiro, Gibbs, Files, Hager, Shehu, Prekker, Erickson, Exline, Gong, Mohamed, Johnson, Srinivasan, Steingrub, Peltan, Brown, Martin, Monto, Khan, Hough, Busse, ten Lohuis, Duggal, Wilson, Gordon, Qadir, Chang, Mallow, Rivas, Babcock, Kwon, Halasa, Grijalva, Rice, Stubblefield, Baughman, Womack, Rhoads, Self. Statistical analysis and data management: Tenforde, Lindsell, Hart, Zhu, Adams, Olson. Funding acquisition: Self. Critical review of the manuscript for important intellectual content: Lauring, Tenforde, Chappell, Gaglani, Ginde, McNeal, Ghamande, Douin, Talbot, Casey, Mohr, Zepeski, Shapiro, Gibbs, Files, Hager, Shehu, Prekker, Erickson, Exline, Gong, Mohamed, Johnson, Srinivasan, Steingrub, Peltan, Brown, Martin, Monto, Khan, Hough, Busse, ten Lohuis, Duggal, Wilson, Gordon, Qadir, Chang, Mallow, Rivas, Babcock, Kwon, Halasa, Grijalva, Rice, Stubblefield, Baughman, Womack, Rhoads, Lindsell, Hart, Zhu, Adams, Schrag, Olson, Kobayashi, Verani, Patel, Self. The corresponding author (Dr. Self) attests that all listed authors meet authorship criteria and that no others meeting the criteria have been omitted.

### Patient Consent

not applicable.

### Ethical Approval

This program was approved as a public health surveillance activity with waiver of informed consent by institutional review boards at the US Centers for Disease Control and Prevention (CDC), the program’s coordinating center at Vanderbilt University Medical Center and each participating site.

### Data Sharing

No additional data are available.

## Data Availability

Additional data produced in this work are not available.

