## Supplementary Materials for "Clinical Severity and mRNA Vaccine Effectiveness for Omicron, Delta, and Alpha SARS-CoV-2 Variants in the United States: A Prospective Observational Study"

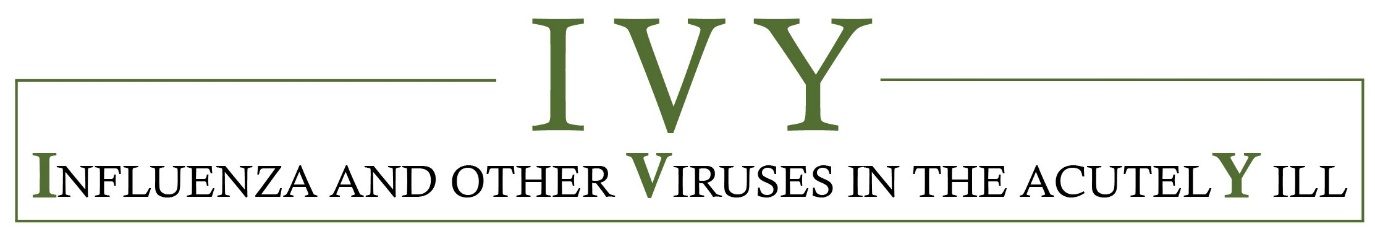

**Supplementary Material**

**Clinical Severity and mRNA Vaccine Effectiveness for Omicron, Delta, and Alpha SARS-CoV-2 Variants in the United States: A Prospective Observational Study**

**The IVY Network**

**Corresponding Author:** Wesley H. Self, MD, MPH; Vanderbilt University Medical Center; 312 Oxford House, 1313 21^st^ Avenue South, Nashville, Tennessee 37232.; phone: 615-936-8047; fax: 615-936-3754.

**Funding:** Primary funding for this study was provided by the United States Centers for Disease Control and Prevention (75D30121F00002).

**Disclaimer:** The findings and conclusions in this report are those of the authors and do not necessarily represent the official position of the Centers for Disease Control and Prevention.

**Contents of Supplementary Materials**

### **I. Supplementary Appendix A.** Investigators and Collaborators

This appendix displays the investigators and collaborators in the Influenza and Other Viruses in the Acutely Ill (IVY) Network by site.

**Baylor, Scott, and White, Temple, Texas**

Manjusha Gaglani, Tresa McNeal, Shekhar Ghamande, Nicole Calhoun, Kempapura Murthy, Judy Herrick, Amanda McKillop, Eric Hoffman, Martha Zayed, Michael Smith

**Baystate Medical Center, Springfield, Massachusetts**

Jay Steingrub, Ryan Kindle, Lori-Ann Kozikowski, Lesley De Souza, Scott Ouellette, Sherell Thornton-Thompson

**Beth Israel Medical Center, Boston, Massachusetts**

Nathan I. Shapiro, Michael Bolstad, Brianna Coviello, Robert Ciottone, Arnaldo Devilla, Ana Grafals, Conor Higgins, Lisa Kurt, Carlo Ottanelli, Kimberly Redman, Douglas Scaffidi, Alexander Weingart

**Centers for Disease Control and Prevention (CDC), Atlanta, Georgia**

Miwako Kobayashi, Samantha Olson, Manish Patel, Mark Tenforde, Meagan Stephenson, Stephanie Schrag, Jennifer Verani, Katherine Adams

**Cleveland Clinic, Cleveland, Ohio**

Abhijit Duggal, Omar Mehkri, Meg Mitchell, Connery Brennan, Kiran Ashok, Bryan Poynter

**Emory University, Atlanta, Georgia**

Laurence Busse, Caitlin ten Lohuis, Nicholas Stanley

**Hennepin County Medical Center, Minneapolis, Minnesota**

Matthew Prekker, Heidi Erickson, Audrey Hendrickson, Sean Caspers, Walker Tordsen, Olivia Kaus, Ellen Maruggi, Tyler Scharber

**Intermountain Medical Center, Murray, Utah**

Ithan Peltan, Samuel Brown, Jeffrey Jorgensen, Robert Bowers, Jennifer King, Valerie Aston

**Johns Hopkins University, Baltimore, Maryland**

David N, Hager, Arber Shehu, Richard E. Rothman

**Montefiore Medical Center, Bronx, New York**

Michelle Gong, Amira Mohamed, Rahul Nair, Jen-Ting (Tina) Chen

**Ohio State Medical Center, Columbus, Ohio**

Matthew Exline, Sarah Karow, Emily Robart, Paulo Nunes Maldonado, Maryiam Khan, Preston So

**Oregon Health and Sciences University, Portland, Oregon**

Akram Khan, Catherine L. Hough, Olivia Krol, Zachary Zouyed, Michael Acosta, Reihaneh Bazyarboroujeni

**Stanford University, Stanford, California**

Jennifer G. Wilson, Alexandra June Gordon, Joe Levitt, Cynthia Perez, Anita Visweswaran, Jonasel Roque

**University of California-Los Angeles, Los Angeles, California**

Nida Qadir, Steven Chang, Adreanne Rivera, Trevor Frankel

**University of Colorado, Aurora, Colorado**

Adit Ginde, David Douin, Jennifer Goff, David Huynh, Kelly Jensen, Conner Driver, Michael Carricato, Ian Chambers

**University of Iowa, Iowa City, Iowa**

Nick Mohr, Anne Zepeski, Paul Nassar, Lori Stout, Zita Sibenaller, Alicia Walter, Jasmine Mares, Logan Olson, Bradley Clinansmith

**University of Miami, Miami, Florida**

Chris Mallow, Hayley Gershengorn, Carolina Rivas

**University of Michigan, Ann Arbor, Michigan**

Emily Martin, Arnold Monto, Adam Lauring, EJ McSpadden, Rachel Truscon, Anne Kaniclides, Lara Thomas, Ramsay Bielak, Weronika Damek Valvano, Rebecca Fong, William J. Fitzsimmons, Christopher Blair, Andrew L. Valesano, Julie Gilbert

**University of Washington, Seattle, Washington**

Daniel J. Henning, Christine D. Crider, Kyle A. Steinbock, Thomas C. Paulson, Layla A. Anderson

**Vanderbilt University Medical Center, Nashville, Tennessee**

Wesley H. Self, H. Keipp Talbot, Chris Lindsell, Carlos Grijalva, Ian Jones, Natasha Halasa, James Chappell, Kelsey Womack, Jillian Rhoads, Adrienne Baughman, Christy Kampe, Jakea Johnson, Kim Hart, Robert McClellan, Todd Rice, Jonathan Casey, William B. Stubblefield, Yuwei Zhu, Laura L. Short, Lauren J. Ezzell, Margaret E. Whitsett, Rendie E. McHenry, Samarian J. Hargrave, Marica Blair, Jennifer L. Luther, Claudia Guevara Pulido, Bryan P. M. Peterson

**Wake Forest University, Winston-Salem, North Carolina**

D. Clark Files, Kevin Gibbs, Mary LaRose, Leigha Landreth, Madeline Hicks, Lisa Parks

**Washington University, St. Louis, Missouri**

Hilary Babcock, Jennie Kwon, Jahnavi Bongu, David McDonald, Candice Cass, Sondra Seiler, David Park, Tiffany Hink, Meghan Wallace, Carey-Ann Burnham, Olivia G. Arter

### **II. Supplementary Appendix B.** Supplemental Methods

1. **Enrollment practices**

Adults admitted to 21 hospitals in the United States were enrolled into this study. Three cohorts of hospitalized patients were enrolled: COVID-19 cases, test-negative controls, and syndrome negative controls. Eligibility criteria for each cohort are detailed in the next section. In brief, cases had symptoms consistent with COVID-19 and a positive test for SARS-CoV-2. Test negative controls had symptoms potentially consistent with COVID-19 but had a negative test for SARS-CoV-2. Syndrome negative controls did not have symptoms consistent for COVID-19, were hospitalized for a reason other than acute respiratory illness and had a negative test for SARS-CoV-2. Enrollment of syndrome negative controls ended in October 2021 based on the findings that vaccine effectiveness results were nearly identical when separately using test negative controls and syndrome negative controls; the data demonstrated that test negative controls alone provided an adequate control group alone. After October 2021, enrollment of cases and test negative controls continued.

Patients were enrolled as cases and controls based on the results of clinical SARS-CoV-2 testing at the local hospital laboratory. Respiratory samples were also collected by study personnel and shipped to Vanderbilt University Medical Center for independent SARS-CoV-2 RT-PCR testing. Final case/control classification was based SARS-CoV-2 test results from both the local clinical laboratories and the central laboratory. Patients enrolled as a case or test negative control who had any positive SARS-CoV-2 test (either a clinical test or central laboratory test) within 10 days of symptom onset were classified as cases. Patients enrolled as a test negative control or syndrome negative control who had all SARS-CoV-2 tests return negative (both clinical tests and central laboratory tests) were included in the analysis as controls.

During the enrollment period, study personnel screened for eligible cases daily with the intent of enrolling all eligible cases. Study personnel also screened for eligible controls daily, with the intent of enrolling 1 control patient within 14 days of each case patient enrolled. If multiple potential control patients were eligible at the same time, study personnel randomly selected which patient to enroll as a control. As a public health surveillance project, written informed consent was not obtained for participation. A patient was considered enrolled in the surveillance program at the time that the first data were collected from the participant by study personnel.

2. **Eligibility criteria**

The section details the eligibility criteria for enrollment for the 3 cohorts of patients included in the study: COVID-19 cases, test-negative controls, and syndrome negative controls.

***Covid-19 Cases***

Summary for Cohort 1: Adult admitted to the hospital for acute Covid-19 who has tested positive for SARS-CoV-2.

Inclusion for Cohort 1 (cases):

1. Age ≥18 years old.
2. Hospital admission or in an emergency department awaiting hospital admission.
3. Symptoms and/or signs believed to be due to Covid-19, including at least 1 of the following: fever; cough; shortness of breath; loss of taste; loss of smell; use of respiratory support (high flow oxygen by nasal cannula, non-invasive ventilation or invasive ventilation) for the acute illness; new pulmonary findings on chest imaging consistent with pneumonia.
4. Clinically obtained test that is **positive** for acute SARS-CoV-2 infection. The positive test may be obtained before or after hospital arrival. Examples of acute SARS-CoV-2 tests include RT-PCR tests, nucleic acid amplification tests (NAAT), and antigen tests. Serology testing may not be used for eligibility.

Exclusion for Cohort 1 (cases):

1. Previous inclusion as a case.
2. The first positive test for acute SARS-CoV-2 infection is known to have occurred more than 10 days after onset of Covid-19 symptoms/signs listed in inclusion criterion #3. Patients with unknown onset date for Covid-19 symptoms/signs may be included.
3. Hospital presentation for the Covid-19 admission is known to have occurred more than 14 days after onset of Covid-19 symptoms/signs listed in inclusion criterion #3. Patients with unknown onset date for Covid-19 symptoms/signs may be included. Patients transferred from other hospitals may be included; the time of hospital presentation is the time of presentation to the first hospital.

***Test Negative Controls***

Summary for Cohort 2: Adult admitted to the hospital for an acute illness with symptom overlap with Covid-19 who has tested negative for SARS-CoV-2.

Inclusion for Cohort 2 (test negative controls):

1. Age ≥18 years old.
2. Hospital admission or in an emergency department awaiting hospital admission.
3. Symptoms and/or signs that overlap with Covid-19, including at least one of the following: fever; cough; shortness of breath; loss of taste; loss of smell; use of respiratory support (high flow oxygen by nasal cannula, non-invasive ventilation or invasive ventilation) for the acute illness; new pulmonary findings on chest imaging consistent with pneumonia.
4. Clinically obtained test that is **negative** for acute SARS-CoV-2. The negative test may be obtained before or after hospital arrival. Examples of acute SARS-CoV-2 tests include RT-PCR tests, NAAT, and antigen tests. Serology testing may not be used for eligibility.

Exclusion for Cohort 2 (test negative controls):

1. Previous inclusion as a control.
2. The first negative test for acute SARS-CoV-2 infection is known to have occurred more than 10 days after onset of symptoms/signs listed in inclusion criterion #3. Patients with unknown onset date for Covid-19 symptoms/signs may be included.
3. Hospital presentation for the admission is known to have occurred more than 14 days after onset of symptoms/signs listed in inclusion criterion #3. Patients with unknown onset date for symptoms/signs may be included. Patients transferred from other hospitals may be included; the time of hospital presentation is the time of presentation to the first hospital.
4. Any positive test for acute SARS-CoV-2 infection in the 14 days prior to hospital presentation or between hospital presentation and inclusion (patients with a positive acute SARS-CoV-2 test should be screened for potential inclusion as a case).

***Syndrome-negative controls***

Summary for Cohort 3: Adult admitted to the hospital for a reason other than an acute respiratory illness and who does not have a clinical suspicion for Covid-19.

Inclusion for Cohort 3 (syndrome negative controls):

1. Age ≥18 years old.
2. Hospital admission or in an emergency department awaiting admission.
3. Clinical impression that Covid-19 is not the reason for admission.
4. None of the following signs or symptoms that overlap with Covid-19 in the past 14 days: fever; cough; shortness of breath; loss of taste; loss of smell; use of respiratory support (high flow oxygen by nasal cannula, non-invasive ventilation or invasive ventilation) for the acute illness; new pulmonary findings on chest imaging consistent with pneumonia.

Exclusion for Cohort 3 (syndrome negative controls):

1. Previous inclusion as a control.
2. Any positive test for acute SARS-CoV-2 infection in the 14 days prior to hospital presentation or between hospital presentation and inclusion (patients with a positive acute SARS-CoV-2 test should be screened for potential inclusion as a case).

### **III. Supplementary Appendix C.** Vaccine effectiveness for partial vaccination with an mRNA vaccine.

This section describes results for vaccine effectiveness calculations for partial vaccination with mRNA COVID-19 vaccines, defined as receipt of a single vaccine dose or receipt of 2 vaccine doses with COVID-19 illness onset within 14 days of receipt of the second vaccine dose. A single dose of an mRNA COVID-19 vaccine is not a recommended regimen by the United States (US) Centers for Disease Control and Prevention (CDC) and is not authorized or approved by the US Food and Drug Administration (FDA). Some people receive a first dose of an mRNA vaccine and then do not follow through with obtaining subsequent vaccine doses. Additionally, some individuals may receive a second vaccine dose and develop illness shortly after the second dose is received and before full immune protection associated with the second vaccine dose develops. Thus, we estimated the vaccine effectiveness of partial vaccination with mRNA vaccines. This is a combined analysis for the two mRNA COVID-19 vaccines available in the US -- BNT162b2 (Pfizer-BioNTech) and mRNA-1273 (Moderna). Further, little or no vaccine-associated protection against SARS-CoV-2 is expected over several days immediately following receipt of a first mRNA vaccine dose. We additionally estimated vaccine effectiveness in patients who received 1 mRNA vaccine dose 0-13 days before illness onset as a “bias indicator.”

Similar to the vaccine effectiveness calculation described in the main text for 2 and 3 doses of an mRNA vaccine, vaccine effectiveness for partial vaccination with an mRNA vaccine to prevent COVID-19 hospitalization was calculated by using a test-negative design, in which the odds of antecedent vaccination were compared between cases and controls. Three vaccination groups were considered: Group 1) Patients who received 1 vaccine dose 0-13 days before illness onset; Group 2) patients who received 1 vaccine dose ≥14 days before illness onset; and Group 3) patients who received 2 vaccine doses, with the first dose received ≥14 days before illness onset and the second dose received 0-13 days before onset. A multivariable unconditional logistic regression model was constructed with case-control status as the dependent variable, vaccination status (vaccinated vs. unvaccinated) as the primary independent variable and the following covariables selected *a priori*: calendar date of admission in biweekly intervals, US Department of Health and Human Services region (10 regions), age, sex, and self-reported race and Hispanic ethnicity. Vaccine effectiveness to prevent COVID-19 hospitalization [VE(hospitalization)] was calculated with the adjusted odds ratio (aOR) from this model as: VE(hospitalization) = (1 – aOR) × 100. For this one-dose vaccine effectiveness calculation, we combined all time periods (Alpha, Delta, and Omicron) together.

Overall, 255 vaccinated patients were included in Group 1, 493 in Group 2, and 185 in Group 3. The time between a receipt of the first vaccine dose and illness onset was short, with a median of 6 (IQR 3 to 9) days, 48 (IQR 21-132) days, and 32 (IQR 28-38) days between receipt of the first vaccine dose and the date of illness onset in groups 1, 2, and 3, respectively. The time interval is short because most people in the US who obtained one mRNA dose also received a second dose in the subsequent weeks. Thus, the vaccine effectiveness estimates reported here are generally limited to only the first several weeks after receipt of the vaccine. Waning effectiveness from a single mRNA vaccine dose is expected over time but could not be rigorously measured in this study due to the small number of patients with extended periods of time since a single vaccine dose.

Estimated vaccine effectiveness for Group 1 (single vaccine dose 0-13 days prior to illness onset) was 16% (95% CI: -10 to 36%), for Group 2 (single vaccine dose ≥14 days prior to illness onset) was 77% (95% CI: 71 to 81%), and for Group 3 (2 vaccine doses with second vaccine dose 0-13 prior to illness onset) was 84% (95% CI: 74 to 89%). Combining Groups 2 and Group 3 as “partially vaccinated” resulted in vaccine effectiveness of 79% (95% CI: 74 to 82%).

### **IV. Supplementary Tables**

#### Table S1. COVID-19 vaccine effectiveness (VE) publications from the IVY Network.

The IVY Network publishes VE estimates iteratively, with later publications adding sample size and focusing on specific questions that were not addressed in earlier publications. Some participants are included in multiple publications. The current manuscript includes participants enrolled between March 11, 2021, and January 14, 2022 with a focus on variant-specific VE and severity.

| **Publication** | **Primary Question Addressed** | **Data cut**  **(cohort initiated March 11, 2021)** | **Sample size** | **Primary finding** |
| --- | --- | --- | --- | --- |
| Clin Infect Dis 2021. PMID: 34358310. | COVID-19 mRNA VE against hospitalization during the early phase of the US COVID-19 program. | May 5, 2021 | 1,212 | mRNA VE against COVID-19 hospitalization among US adults = 87.1% (95% CI: 80.7 – 91.3%) |
| MMWR 2021; 70:1156. PMID: 34437524 | COVID-19 mRNA VE against hospitalization beyond 12 weeks from full vaccination. | July 14, 2021 | 3,089 | mRNA VE against COVID-19 hospitalization from 13 to 24 weeks post-vaccination among US adults = 84% (95% CI: 77 – 90%) |
| MMWR 2021; 70:1337. PMID: 34555004 | Comparative effectiveness of COVID-19 vaccines available in the US for preventing COVID-19 hospitalizations among immunocompetent adults | August 15, 2021 | 3,689 | VE against COVID-19 hospitalizations varied by vaccine product among immunocompetent adults – Moderna: 93% (95% CI: 91-95%); Pfizer BioNTech: 88% (95% CI: 85-91%); Janssen: 71% (95% CI: 56-81%). |
| JAMA 2021.  PMID: 34734975 | Effectiveness of mRNA vaccines to prevent disease progression to critical illness or death among adults hospitalized with COVID-19. | August 15, 2021 | 4,515 | Prior vaccination was associated with a 67% (95% CI: 42-81%) relative risk reduction lower risk of progression to invasive mechanical ventilation or death among adults hospitalized with COVID-19, suggesting vaccination attenuated disease severity. |
| Current manuscript | Clinical Severity and mRNA Vaccine Effectiveness for Omicron, Delta, and Alpha SARS-CoV-2 Variants in the United States: A Prospective Observational Study | January 14, 2022 | 11,690 | VE against COVID-19 during the period that Delta variant dominated was 85% (95% CI: 83 to 87%) for 2 mRNA vaccine doses and 94% (95% CI: 92 to 95%) for 3 vaccine doses; for the early period that Omicron variant dominated VE was 65% (95% CI: 51 to 75%) for 2 doses and 86% (95% CI: 77 to 91%) for 3 doses. |

#### Table S2. Modified World Health Organization COVID-19 Clinical Progression Scale.

This scale was used in this analysis to assess disease severity among adults hospitalized with COVID-19.

| Patient State | Descriptor | Severity Level |
| --- | --- | --- |
| Uninfected | Uninfected; no viral RNA detected | 0  [not studied in this analysis] |
| Ambulatory mild disease | Asymptomatic; viral RNA detected | 1  [not studied in this analysis] |
|  | Symptomatic; independent | 2  [not studied in this analysis] |
|  | Symptomatic; assistance needed | 3  [not studied in this analysis] |
| Hospitalized: moderate disease | Hospitalized; no oxygen therapy | 4 |
|  | Hospitalized; standard oxygen therapy by mask or nasal prongs | 5 |
| Hospitalized: severe disease | Hospitalized, oxygen by high flow nasal cannula or non-invasive ventilation | 6 |
|  | Invasive mechanical ventilation | 7 |
|  | Invasive mechanical ventilation plus other organ support including ECMO, vasopressors or new renal replacement therapy | 8 |
| Death | Death | 9 |

ECMO = extracorporeal membrane oxygenation; HFNC = high-flow nasal cannula; NIV = non-invasive ventilation

^a^Uninfected and mild severity were not included in this analysis that was restricted to hospitalized, laboratory-confirmed COVID-19 patients.

#### Table S3. SARS CoV-2 variants identified by sequencing.

This table displays the SARS-CoV-2 variants identified by viral whole genome sequencing during the Alpha period (March 11 – July 3, 2021), Delta period (July 4, 2021 – December 25, 2021), and Omicron period (December 26, 2021 – January 14, 2022).

| **SARS-CoV-2 Variant** | **Sequenced Cases during Alpha Period (March 11 – July 3, 2021)**  **[n = 421]** | **Sequenced Cases during Delta Period (July 4 – December 25, 2021)**  **[n = 1930]** | **Sequenced Cases during Omicron Period (December 26, 2021 – January 14, 2022)**  **[n = 248]** |
| --- | --- | --- | --- |
| Alpha | 242 (57.5%) | 5 (0.3%) | 0 (0%) |
| Delta | 46 (10.9%) | 1867 (96.7%) | 58 (23.4%) |
| Beta | 6 (1.4%) | 0 (0%) | 0 (0%) |
| Gamma | 38 (9.0%) | 6 (0.3%) | 0 (0%) |
| Omicron | 0 (0%) | 37 (1.9%) | 190 (76.6%) |
| Other variant* | 89 (21.1%) | 15 (0.8%) | 0 (0%) |

*Other variants included: B.1.526 (14), B.1.621 (12), B.1.526.1 (11), B.1.429 (11), B.1 (10), B.1.1.519 (7), B.1.2 (7), B.1.526.3 (4), B.1.621.1 (4), B.1.1.28 (3), B.1.526.2 (3), C.37 (3), B.1.525 (2), B.1.623 (2), B.1.628 (2), B.1.1.318 (1), B.1.1.372 (1), B.1.361 (1), B.1.441 (1), B.1.517 (1), B.1.612 (1), B.1.637 (1), C.36.3 (1), R.1 (1).

#### Table S4. Patient characteristics of by sequence-confirmed cases.

This table displays patient characteristics of controls and COVID-19 cases with sequence-confirmed Alpha, Delta, and Omicron variant COVID-19 include in vaccine effectiveness analyses. Alternatively, baseline characteristics based on period of enrollment (Alpha, Delta, and Omicron periods) are shown in Table 1.

| **Patient Characteristic** | **All controls**  **(n=5962)** | **Sequenced Alpha cases**  **(n=247)** | **Sequenced Delta cases**  **(n=1971)** | **Sequenced Omicron cases**  **(n=227)** |
| --- | --- | --- | --- | --- |
| Age in years, median (IQR) | 63 (50-72) | 60 (48-68) | 60 (47-71) | 61 (48-71) |
| Female sex, No. (%) | 2975 (49.9) | 115 (46.6) | 886 (45.0) | 114 (50.2) |
| Race and ethnicity, No. (%) |  |  |  |  |
| Non-Hispanic White | 3611 (60.6) | 123 (49.8) | 1071 (54.3) | 80 (35.2) |
| Non-Hispanic Black | 1240 (20.8) | 70 (28.3) | 413 (21.0) | 72 (31.7) |
| Hispanic, any race | 772 (12.9) | 45 (18.2) | 346 (17.6) | 55 (24.2) |
| Non-Hispanic, Other | 253 (4.2) | 8 (3.2) | 98 (5.0) | 18 (7.9) |
| Unknown | 86 (1.4) | 1 (0.4) | 43 (2.2) | 2 (0.9) |
| US Census region, No. (%) |  |  |  |  |
| Northeast | 885 (14.8) | 26 (10.5) | 383 (19.4) | 47 (20.7) |
| South | 2371 (39.8) | 80 (32.4) | 628 (31.9) | 98 (43.2) |
| Midwest | 1374 (23.0) | 91 (36.8) | 519 (26.3) | 48 (21.1) |
| West | 1332 (22.3) | 50 (20.2) | 441 (22.4) | 34 (15.0) |
| Resident of long-term care facility, No. / Total (%) | 321/5778 (5.6) | 4/245 (1.6) | 74/1867 (4.0) | 14/216 (6.5) |
| ≥1 prior hospitalization in past year, No. / Total (%) | 3031/5537 (54.7) | 65/215 (30.2) | 522/1778 (29.4) | 109/224 (48.7) |
| Current tobacco use, No. / Total (%) | 1016/5302 (19.2) | 21/200 (10.5) | 173/1650 (10.5) | 31/202 (15.3) |
| Number of chronic medical conditions‡, median (IQR) | 2 (1-3) | 2 (1-3) | 2 (1-3) | 2 (1-3) |
| Categories of medical conditions‡ |  |  |  |  |
| Chronic cardiovascular disease | 4158 (69.7) | 137 (55.5) | 1164 (59.1) | 150 (66.1) |
| Chronic pulmonary disease | 1973 (33.1) | 65 (26.3) | 429 (21.8) | 63 (27.8) |
| Diabetes mellitus | 1962 (32.9) | 75 (30.4) | 645 (32.7) | 75 (33.0) |
| Immunocompromising condition | 1458 (24.5) | 46 (18.6) | 375 (19.0) | 77 (33.9) |
| Obesity, No. / Total (%) | 2391/5900 (40.5) | 146/243 (60.1) | 1022/1949 (52.4) | 97/225 (43.1) |
| Vaccination status |  |  |  |  |
| Unvaccinated | 2054 (34.5) | 226 (91.5) | 1362 (69.1) | 92 (40.5) |
| 2 doses (<150 days) | 2029 (34.0) | 21 (8.5) | 188 (9.5) | 18 (7.9) |
| 2 doses (≥150 days) | 1411 (23.7) | 0 (0) | 380 (19.3) | 77 (33.9) |
| 3 doses | 468 (7.8) | 0 (0) | 41 (2.1) | 40 (17.6) |
| If vaccinated, vaccine product received, No. / Total (%) |  |  |  |  |
| **Patient Characteristic** | **All controls**  **(n=5962)** | **Sequenced Alpha cases**  **(n=247)** | **Sequenced Delta cases**  **(n=1971)** | **Sequenced Omicron cases**  **(n=227)** |
| BNT162b2 (Pfizer-BioNTech) | 2269/3908 (58.1) | 13/21 (61.9) | 395/609 (64.9) | 92/135 (68.1) |
| mRNA-1273 (Moderna) | 1615/3908 (41.3) | 7/21 (33.3) | 210/609 (34.5) | 41/135 (30.4) |
| Mixed products | 24/3908 (0.6) | 1/21 (4.8) | 4 (0.7) | 2 (1.5) |
| Days since dose 3 if 3 doses received, median (IQR) | 41 (23-64) | --- | 38 (21-71) | 69.5 (36-107.5) |

Definitions: IQR = interquartile range; US = United States

‡ Chronic medical conditions were obtained from structured medical chart review and body-mass index calculated using documented height and weight.

### **V. Supplementary Figures**

#### Figure S1. Flow diagram of participant participation.

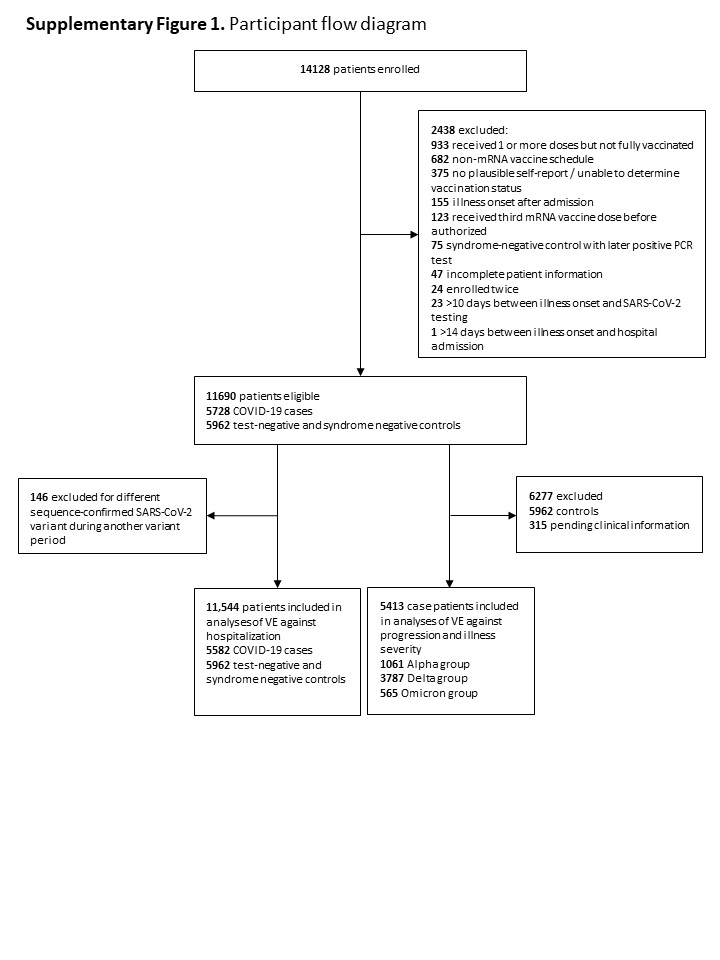
